# The prevalence of type 2 diabetes mellitus among people with amyotrophic lateral sclerosis is increasing in the United States

**DOI:** 10.64898/2026.10.27.26366623

**Authors:** Xing Song, Jaime Raymond, Paul Mehta, Ikjae Lee, The Greater Plain Collaborative

## Abstract

**Background:** Type 2 diabetes mellitus (T2DM) has been associated with lower risk of ALS and prevalence of comorbid T2DM has been reported to be lower among people with ALS (PALS) compared to general population. We examined the trend of comorbid DM prevalence among PALS in comparison to overall population from 2011 to 2021 in the United States (US).

**Methods:** We used multi-center electronic health records (EHRs) data within the GPC Reusable Observable Unified Study Environment (GROUSE) and validated algorithms to identify PALS with T2DM. Prevalence was defined by number of cases per population in each year. The trend of changes in prevalences from 2011 to 2021 was examined using mixed linear models, adjusted for age and sex.

**Results:** A total of 25,676,029 unique persons included in the EHRs from 2011 to 2021 were identified. Among them, 1,720,524 (6.7%) had T2DM, 12,388 (0.05%) had ALS, and 1,498 (0.006%) had both ALS/T2DM. Among PALS, prevalence of T2DM increased from 5.2% in 2011 to 12.1% in 2021, 132.7% relative increase in this period. In the overall population, T2DM prevalence increased from 5.9% in 2011 to 7.6% in 2021, a relative increase of 29% in the same period. In the overall population, ALS prevalence increased from 30.2 per 100,000 in 2011 to 33.0 per 100,000 in 2021, a 9.3% increase. In contrast, ALS prevalence increased dramatically among T2DM by 118.7% from 14.4 per 100,000 in 2011 to 31.5 per 100,000 in

2021.

**Discussion:** Prevalence of ALS among T2DM have increased significantly over the past decade, out pacing the trend among general population. Etiologies behind this trend need to be further investigated.

## Introduction

Amyotrophic lateral sclerosis (ALS) is a motor neuron disease with progressive muscle weakness, respiratory failure, and eventual death in 2-5 years from symptom onset.^1^ Previous studies suggested type 2 diabetes mellitus (T2DM) is associated with a lower risk of ALS, and the prevalence of T2DM among people with ALS (PALS) is lower than that among population.^2–4^ Surprisingly, the literature is scarce and recent epidemiological studies are lacking.

Diabetes mellitus (DM) is a group of metabolic disorders characterized by dysregulated blood glucose levels.^5^ Type 1 DM (T1DM) is characterized by low or absent insulin secretion from the pancreas, and treatment is directed toward providing insulin. In comparison, T2DM involves tissue resistance to insulin, resulting in initially high levels of insulin production for compensation which eventually decreases. Treatment of T2DM involves several classes of drugs with different mechanisms: providing insulin; reducing insulin resistance; reducing hepatic output of glucose; suppressing glucose intake and absorption.^6^

T2DM treatment landscape evolved over the last 2 decades. Metformin was introduced to US in 1995 and has been established as the first line treatment.^7^ It lowers blood glucose levels through reducing hepatic glucose output and increase peripheral glucose utilization. Glucagon-like peptide 1 receptor agonists (GLP-1RA) are a new class of drugs introduced in 2005 that prolong the activation of GLP-1 receptors, leading to delayed gastric emptying, decreased blood glycemic responses, and decreased appetite. GLP-1RAs are increasingly used for T2DM and obesity for their weight loss effect.^8^ While weight loss and low adiposity are associated with cardiovascular benefits in the general population,^9^ they have been linked to increased risk of ALS and rapid disease progression after symptom onset.^10–12^ In this study, we examined the trends of ALS, T2DM and ALS/T2DM prevalences among the population and prescription patterns of DM medications from 2011 to 2021.

## Methods

### Data Source

This retrospective study was conducted using data from the Greater Plain Collaborative (GPC) Reusable Observable Unified Study Environment (GROUSE) database, a multicenter real-world data repository that integrates longitudinal electronic health records (EHRs) from 14 participating healthcare systems across the United States.^13^ The GROUSE database leverages the PCORnet Common Data Model (CDM) that harmonizes source EHR, social security death master file, and billing system data into an interoperable structure that enable standardized analyses across institutions while preserving consistent definitions for diagnoses, medications, laboratory results, procedures, and demographic characteristics.^14^

The database contains longitudinal patient-level information, including demographic characteristics, encounter history, diagnoses, laboratory test results, medication prescriptions and dispensing records, procedures, vital status, and healthcare utilization. Data underwent standardized extraction, transformation, and quality assurance procedures before inclusion in the research database. Because the database captures routine clinical care across multiple healthcare settings, it provides relatively comprehensive longitudinal follow-up suitable for estimating disease prevalence, incidence, and medication utilization.

### Study Population

The source population consisted of all individuals receiving care within participating healthcare systems during the study period from 2011-01-01 to 2021-12-31. Patients contributed to annual analyses if they had evidence of active observation during the study period (i.e., with at least one in-person or telehealth visit). Patients were followed longitudinally from study entry until death, loss to follow-up, or the end of the observation period. Demographic characteristics were determined using the most recent available information prior to or during each study year.

### Diabetes Mellitus Case Ascertainment

DM was identified using the validated computable phenotype developed by the SUrveillance, PREvention, and ManagEment of Diabetes Mellitus (SUPREME-DM) DataLink.^15^ This phenotype has been extensively evaluated and applied to identify individuals with diabetes in large, heterogeneous electronic health record–based populations.^15^ The SUPREME-DM phenotype integrates multiple complementary sources of structured clinical information rather than relying exclusively on diagnosis codes. Individuals were classified as having diabetes based on qualifying evidence from one or more of the following domains:

- Diagnosis codes indicative of diabetes, defined by using International Classification of Diseases, Ninth Revision, Clinical Modification (ICD-9-CM), or Tenth Revision, Clinical Modification (ICD-10-CM), codes (Supplementary table 1);
- Laboratory evidence consistent with DM, including elevated fasting plasma glucose or glycated hemoglobin (HbA1c) values that met established diagnostic thresholds; and
- Prescription or medication-dispensing records for insulin or other glucose-lowering medications.

To distinguish DM subtypes, we evaluated two computable phenotypes for identifying type 1 DM (T1DM). The primary approach was based on the All of Us (AoU) Research Program algorithm, which identifies T1DM primarily using diagnosis-code evidence.^16^ The alternative approach was the Klompas algorithm,^17^ which incorporates the relative frequency of T1DM- and type 2 DM (T2DM)-specific diagnosis codes, together with other available clinical information, to differentiate between the two major diabetes subtypes. Primary results were reported using the AoU definition of T1DM. Sensitivity analyses were conducted using the Klompas phenotype to assess the robustness of the findings to alternative diabetes-subtype classification methods.

Individuals who met the overall SUPREME-DM definition for diabetes but did not meet the applicable criteria for T1DM were classified as having T2DM or another non-T1DM diabetes category, as appropriate for the specific analysis.

### ALS Case Ascertainment

ALS cases were identified using an adjusted version of capture–recapture surveillance algorithm developed by the U.S. Centers for Disease Control and Prevention (CDC) for the National ALS Registry.^18,19^ This algorithm was designed to maximize case ascertainment in large healthcare databases by integrating multiple longitudinal sources of clinical information. The algorithm evaluates longitudinal diagnosis histories, healthcare utilization patterns, provider specialties, and ALS-specific clinical characteristics to classify patients according to the probability of having ALS. Patients classified as confirmed ALS cases according to the CDC algorithm were included as ALS cases in this study. Several components of the original CDC algorithm could not be applied consistently because the completeness and standardization of facility type, provider specialty, and Medicare entitlement information varied across participating electronic health record systems. We therefore adapted the algorithm to emphasize clinical elements that were more consistently available across sites, primarily longitudinal ALS diagnosis-code patterns and documented use of ALS-specific medications. Greater weight was given to repeated ALS diagnoses recorded on distinct dates and across separate encounters than to a single isolated diagnosis code. The modified phenotype retained the central principle of the CDC surveillance approach: ALS classification should be based on converging evidence accumulated over time rather than on the presence of a single diagnosis code. This adaptation was intended to preserve high specificity while supporting reproducible implementation across heterogeneous EHR-based data sources.

### Annual Disease Prevalence

Annual prevalences for ALS, T2DM and ALS/T2DM (with both conditions) were estimated for each calendar year. For a given year, the denominator included individuals who were under active observation for any portion of that calendar year, as determined from documented observation start and censoring dates, and who had available age and sex information. Individuals did not contribute to years before their observation start date or after their censoring date. Under the primary carry-forward specification, a condition was considered prevalent beginning on the date of the first observed qualifying diagnosis and was carried forward through the end of the individual’s enrollment or observable follow-up. Thus, once an individual met the case definition for ALS or T2DM, a new diagnosis code was not required in subsequent years for the condition to remain active. A condition contributed to the annual prevalence estimate if its disease-active interval overlapped any portion of the corresponding calendar year.

Annual T2DM prevalence was calculated as the proportion of eligible individuals under observation during each calendar year whose T2DM-active interval overlapped that year. Annual ALS prevalence was similarly estimated in the overall eligible population. To characterize the intersection of the two conditions, we additionally estimated ALS prevalence among individuals with T2DM and T2DM prevalence among individuals with ALS. We also quantified the annual prevalence of concurrent ALS/T2DM, requiring the disease-active intervals for both conditions to overlap the same calendar year. These conditional and joint measures represent annual co-prevalence rather than lifetime comorbidity. For example, a history of T2DM before the observable ALS period would not by itself qualify an individual as having co-prevalent ALS/T2DM unless both conditions were considered active during the same calendar year under the carry-forward definition.

### Annual Prescription Prevalence

Annual prescription prevalence was estimated separately for each calendar year without carrying prescriptions forward across years. The denominator only included patients with evidence of healthcare engagement during the corresponding year, defined as at least one documented in-person or telehealth visit. The numerator included patients in this denominator cohort with at least one prescription for a commonly used antidiabetic medication during the same calendar year. Medication classes included glucagon-like peptide-1 receptor agonists (GLP-1RA), insulin (INS), dipeptidyl peptidase-4 inhibitors (DPP-4), meglitinides (MEG), sodium-glucose cotransporter-2 inhibitors (SGLT2), sulfonylureas (SU), and biguanides (BIGU; primarily metformin). Each patient was counted once per medication class per year, regardless of the number of prescriptions recorded. Prescriptions from prior years were not assumed to remain active or carried forward; thus, annual prescription prevalence reflects the proportion of actively observed patients with documented prescribing of each medication class during that specific year.

### Statistical Analysis

Analyses were restricted to the prespecified age range (18 and older), with age evaluated at the midpoint of each calendar year. Crude prevalence estimates were expressed as proportions or percentages using the appropriate scaling factor. Age- and sex-adjusted prevalence estimates were obtained by direct standardization across joint age-group-by-sex strata. For the primary analysis, stratum weights were derived from the pooled age-sex distribution of the study cohort across all study years, thereby defining a fixed standard population and ensuring comparability of annual estimates over time. Strata not represented in the standard population were assigned a weight of zero. Confidence intervals (CIs) were selected to accommodate the rarity of ALS and avoid the limitations of symmetric Wald intervals. Wilson score intervals were used for crude prevalence proportions. For directly standardized estimates, CIs were calculated using the method of Dobson and colleagues, which maps Poisson confidence limits for the observed event counts onto the directly standardized rate scale.^20^ To characterize changes in demographic composition over time, we additionally summarized annual age, sex, and race distributions separately for the ALS, ALS with T2DM, and ALS without diabetes cohorts. Mean age was reported with its 95% CI, calculated using the standard error of the mean, while the proportions of male and non-White individuals were reported with 95% Wilson score CIs.

## Results

### Study Population and Cohort Characteristics

Between 2011 and 2021, a total of 25,676,029 unique individuals were identified in the GROUSE EHR database. Of these, 1,720,524 (6.7%) had T2DM. A total of 12,388 individuals had confirmed ALS, corresponding to an overall prevalence of approximately 48 per 100,000 individuals. Among PALS, 1,498 had coexisting T2DM, prevalence of 12%. [Table 1] The mean age was 36.2 years (SD, 24.0) in the overall population, compared with 56.5 years (SD, 16.8) among individuals with T2DM and 64.3 years (SD, 12.0) among those with ALS. Individuals with coexisting ALS and T2DM were the oldest subgroup, with a mean age of 66.3 years (SD, 10.8). Compared with individuals with ALS without T2DM or with ALS overall, those with coexisting T2DM were, on average, approximately two years older. Sex distributions also varied across cohorts. Females represented 53.2% of the overall population and 49.8% of individuals with T2DM, whereas males predominated among individuals with ALS (55.6%) and those with both ALS and T2DM (61.4%). The proportion of males was 5.8 percentage points higher in the ALS/T2DM subgroup than in the overall ALS cohort. Differences in racial and ethnic composition were also observed. White individuals accounted for 64.2% of the overall population, 67.8% of individuals with T2DM, 80.1% of individuals with ALS, and 76.2% of those with ALS/T2DM. Black individuals represented 9.1%, 13.4%, 5.0%, and 8.9% of the respective cohorts. Similarly, the proportion of Hispanic/Latino individuals was higher in the ALS/T2DM subgroup (8.1%) than in the overall ALS cohort (4.7%). The proportion of individuals with unknown race or ethnicity varied across cohorts, indicating potential differences in the completeness of demographic documentation.

**Table 1.**
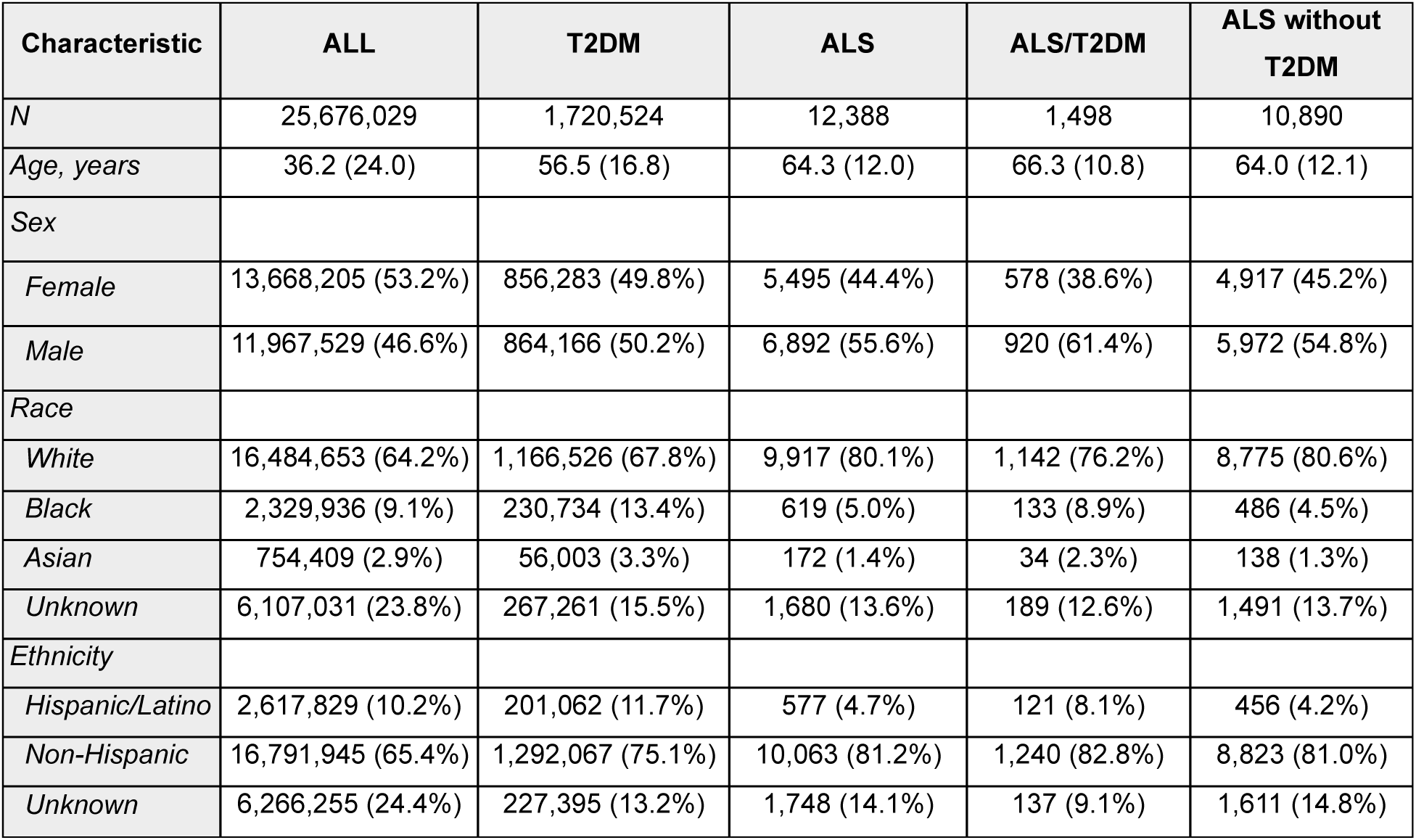
Cohort Characteristics.

| **Characteristic** | **ALL** | **T2DM** | **ALS** | **ALS/T2DM** | **ALS without T2DM** |
| --- | --- | --- | --- | --- | --- |
| *N* | 25,676,029 | 1,720,524 | 12,388 | 1,498 | 10,890 |
| *Age, years* | 36.2 (24.0) | 56.5 (16.8) | 64.3 (12.0) | 66.3 (10.8) | 64.0 (12.1) |
| *Sex* |  |  |  |  |  |
| *Female* | 13,668,205 (53.2%) | 856,283 (49.8%) | 5,495 (44.4%) | 578 (38.6%) | 4,917 (45.2%) |
| *Male* | 11,967,529 (46.6%) | 864,166 (50.2%) | 6,892 (55.6%) | 920 (61.4%) | 5,972 (54.8%) |
| *Race* |  |  |  |  |  |
| *White* | 16,484,653 (64.2%) | 1,166,526 (67.8%) | 9,917 (80.1%) | 1,142 (76.2%) | 8,775 (80.6%) |
| *Black* | 2,329,936 (9.1%) | 230,734 (13.4%) | 619 (5.0%) | 133 (8.9%) | 486 (4.5%) |
| *Asian* | 754,409 (2.9%) | 56,003 (3.3%) | 172 (1.4%) | 34 (2.3%) | 138 (1.3%) |
| *Unknown* | 6,107,031 (23.8%) | 267,261 (15.5%) | 1,680 (13.6%) | 189 (12.6%) | 1,491 (13.7%) |
| *Ethnicity* |  |  |  |  |  |
| *Hispanic/Latino* | 2,617,829 (10.2%) | 201,062 (11.7%) | 577 (4.7%) | 121 (8.1%) | 456 (4.2%) |
| *Non-Hispanic* | 16,791,945 (65.4%) | 1,292,067 (75.1%) | 10,063 (81.2%) | 1,240 (82.8%) | 8,823 (81.0%) |
| *Unknown* | 6,266,255 (24.4%) | 227,395 (13.2%) | 1,748 (14.1%) | 137 (9.1%) | 1,611 (14.8%) |

### Temporal Trends in Diabetes Prevalence by ALS status

The prevalence of T2DM increased over the 11-year study period in both the overall GROUSE EHR population and individuals with ALS [Figure 1]. In the overall population, T2DM prevalence increased from 5.9% in 2011 to 7.6% in 2021, representing an absolute increase of 1.7 percentage points and a relative increase of 28% over this 11-year period. A more pronounced increase was observed among PALS, for whom the crude prevalence of T2DM rose from 5.2% in 2011 to 12.1% in 2021, an absolute increase of 6.9 percentage points and a relative increase of 132.7%. The age-sex-adjusted prevalence consistently showed a significantly higher increase of 5.1 percentage points and a relative increase of 192.5%. In 2011, the age- and sex-adjusted prevalence of T2DM was lower among PALS than in the overall population. However, this difference narrowed over time and was no longer present by 2021 [Figure 1].

**Figure 1.**
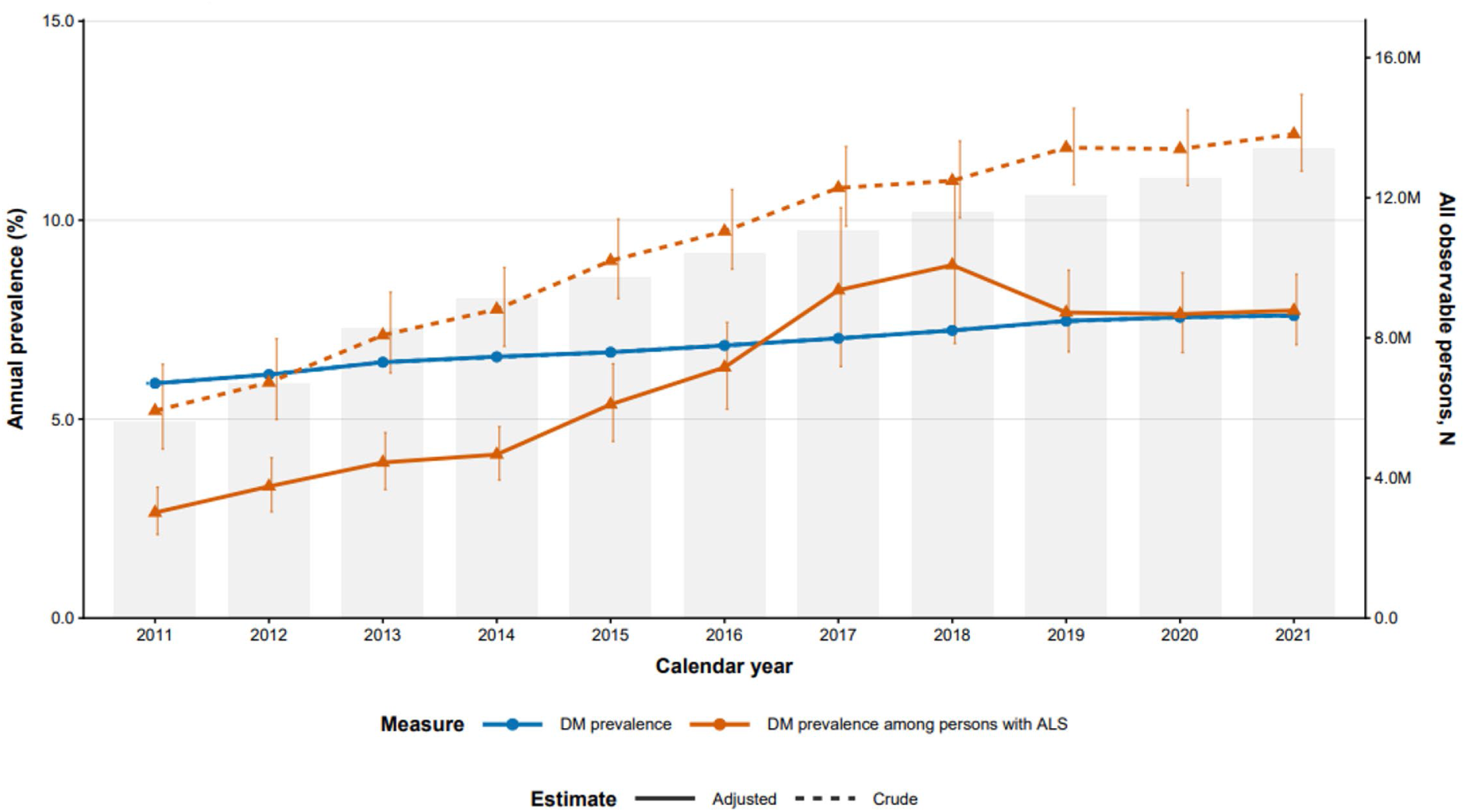
Annual T2DM prevalence in the overall population and among persons with ALS, 2011–2021. Blue, overall population; orange, persons with ALS; solid, age- and sex-adjusted; dashed, crude. Error bars are 95% CIs. Gray bars (right axis) show the number of observable persons per year.

### Temporal Trends in ALS Prevalence by T2DM Status

Temporal trends in ALS prevalence differed by T2DM status [Figure 2]. In the overall population, ALS prevalence remained relatively stable, rising slightly from 30.2 per 100,000 individuals in 2011 to 33.0 per 100,000 in 2021, corresponding to a relative increase of 9.3% over 11 years.

**Figure 2.**
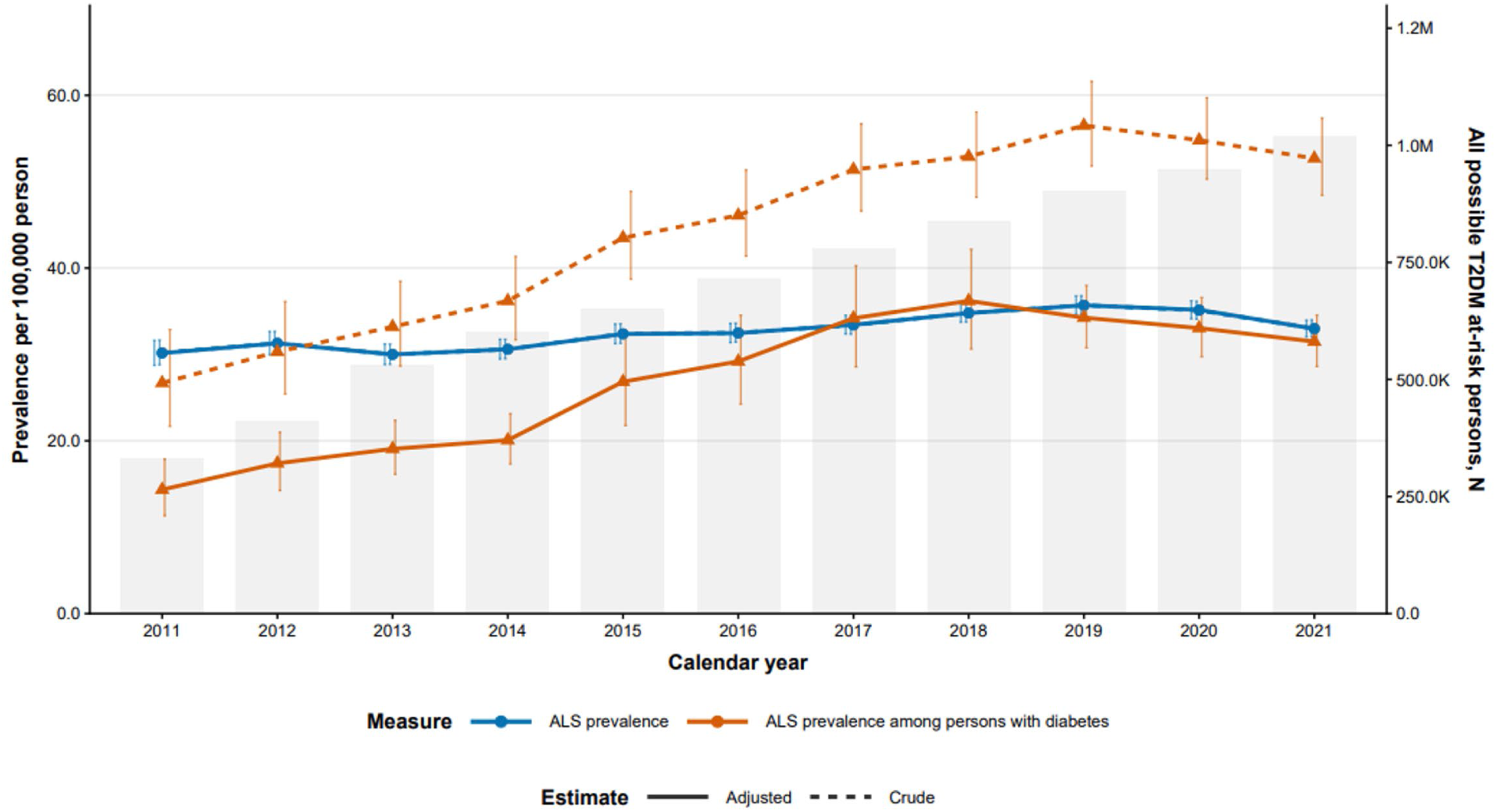
Annual prevalence of amyotrophic lateral sclerosis (ALS) in the overall population and among persons with type 2 diabetes mellitus (T2DM), 2011–2021. Lines show ALS prevalence per 100,000 persons in the overall population (blue) and among persons with T2DM (orange). Solid lines are age- and sex-adjusted estimates, and dashed lines are crude estimates. Error bars represent 95% confidence intervals. Gray bars (right axis) show the number of persons with T2DM at risk in each year.

Among individuals with T2DM, ALS prevalence increased more significantly with an adjusted rate from 14.4 per 100,000 in 2011 to 31.5 per 100,000 in 2021, representing an absolute increase of 17.1 cases per 100,000 and a relative increase of 118.7% in the same period. In 2011, ALS prevalence was lower among individuals with T2DM than among those without T2DM. By 2021, the ALS prevalence among individuals with T2DM is also indifferent from the general ALS prevalence.

### Temporal Trend of Demographic Profiles

Over the 11-year study period, the mean age of PALS increased steadily from 61.9 years (SD, 12.6) in 2011 to 66.1 years (SD, 12.3) in 2021. A similar trend was observed among PALS without T2DM, whose mean age increased from 61.6 years (SD, 12.6) to 65.9 years (SD, 12.5).

[Figure 3] In contrast, PALS with T2DM were consistently older throughout the study period, with mean age remaining relatively stable, ranging approximately from 66 to 69 years (68.6 years in 2011 and 67.7 years in 2021). Consequently, the age gap between PALS with and without T2DM narrowed over time, from 7.0 years in 2011 to 1.8 years in 2021. The proportion of males remained relatively stable among PALS overall (56.0% in 2011 versus 56.4% in 2021) and those without diabetes (55.8% versus 55.6%). In comparison, PALS with T2DM consistently had a higher proportion of males, although this proportion fluctuated over time, from 60.2% in 2011 to 62.4% in 2021. The proportion of non-White individuals declined across all three cohorts, from 30.6% to 24.1% among PALS overall, 30.4% to 23.7% among those without diabetes, and 34.1% to 27.0% among those with T2DM. Despite this overall decline, the proportion of non-White individuals remained consistently higher among PALS with T2DM than among those without diabetes.

**Figure 3.**
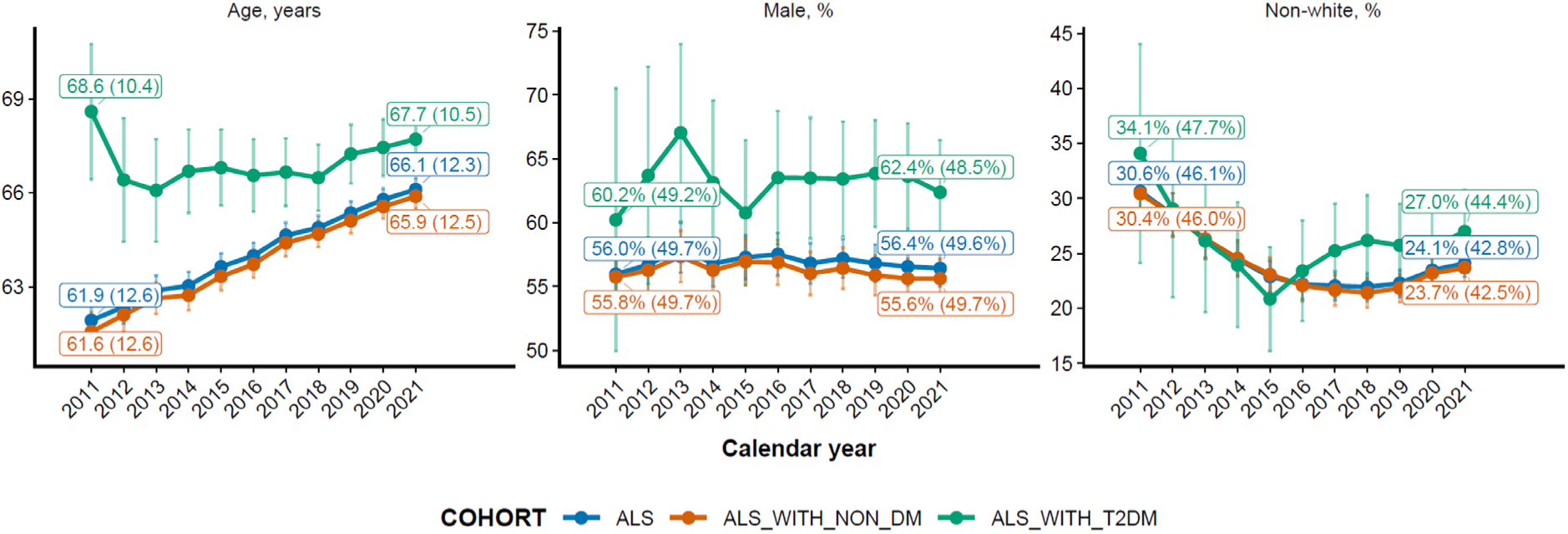
Annual age, sex, and race distribution of persons with ALS by T2DM status, 2011–2021. Mean age (years), % male, and % non-White for all ALS (blue), ALS without T2DM (orange), and ALS with T2DM (green). Error bars are 95% CIs.

### Patterns of Glucose-Lowering Medication Prescriptions

Prescription patterns for glucose-lowering medications over the 11-year study period are presented in Figure 4. Biguanides (BIGU), primarily metformin, remained the most prescribed class of glucose-lowering medications throughout the study period, with prescription prevalence increasing steadily from 52.0% in 2011 to 65.4% in 2021, representing an absolute increase of 13.4 percentage points. GLP-1RAs exhibited the most pronounced relative increase among the medication classes examined, with prescription prevalence rising from 0.7% in 2011 to 19.6% in 2021, corresponding to an absolute increase of 18.9 percentage points and a 28-fold increase relative to baseline. Notably, the upward trajectory of GLP-1RA prescriptions accelerated around 2017, suggesting a shift in prescribing patterns during the latter half of the study period.

**Figure 4.**
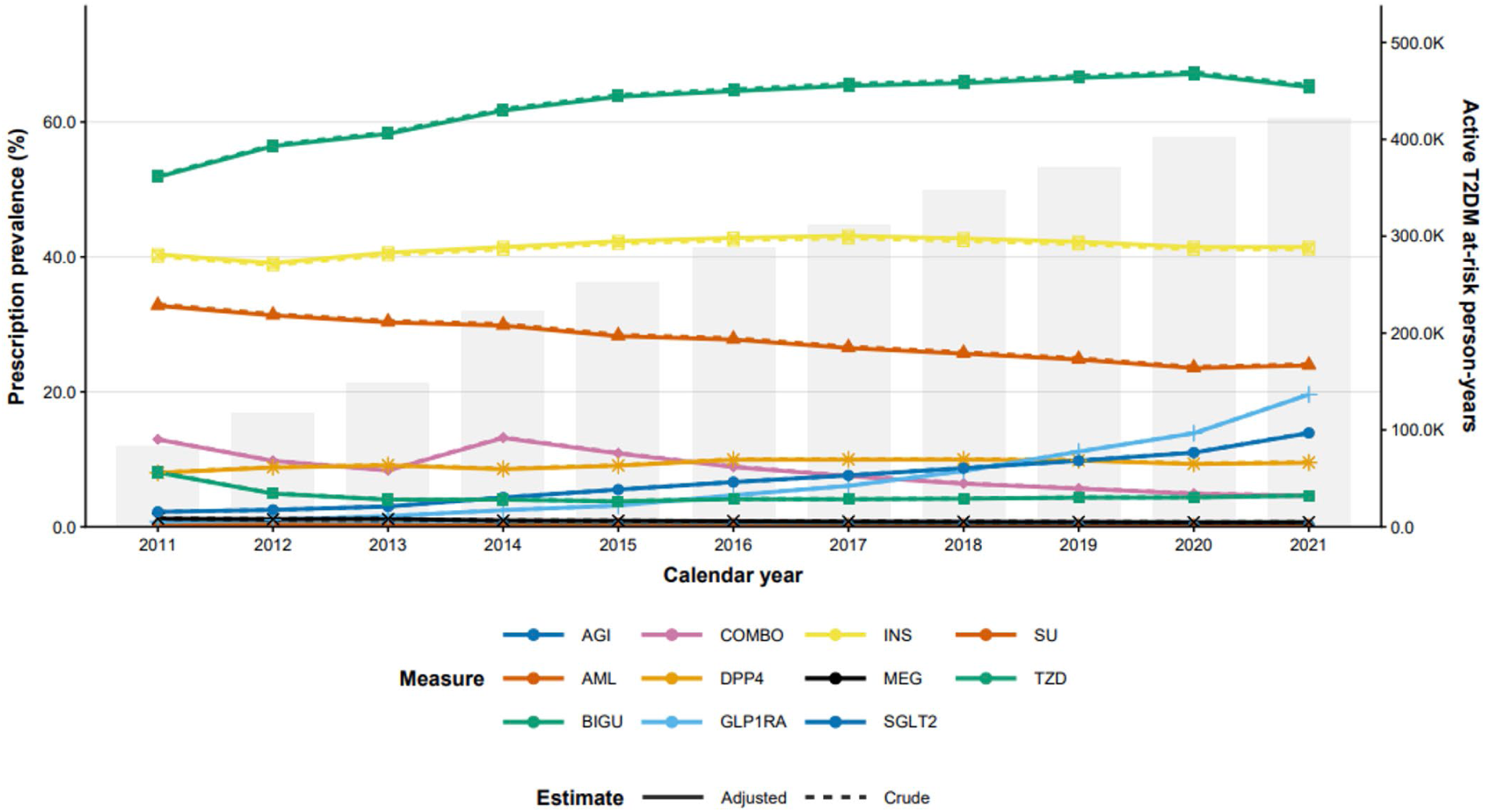
Annual prevalence of glucose-lowering medication prescriptions among persons with type 2 diabetes mellitus (T2DM), 2011–2021. Lines show the percentage of active T2DM person-years with at least one prescription in each medication class. Solid lines are age- and sex-adjusted estimates, and dashed lines are crude estimates. Error bars represent 95% confidence intervals. Gray bars (right axis) show the number of active T2DM at-risk person-years in each year. AGI, alpha-glucosidase inhibitors; AML, amylin analogs; BIGU, biguanides; COMBO, combination products; DPP4, dipeptidyl peptidase-4 inhibitors; GLP1RA, glucagon-like peptide-1 receptor agonists; INS, insulin; MEG, meglitinides; SGLT2, sodium-glucose cotransporter-2 inhibitors; SU, sulfonylureas; TZD, thiazolidinediones.

## Discussion

In this EHR-data-based study of a broad hospital network including more than 25 million individuals actively receiving care in the United States, we observed a substantial increase in the prevalence of T2DM among people with ALS over the past decade. Although T2DM was less common among people with ALS than in the general population at the beginning of the study period, this difference progressively narrowed and disappeared after 2016, accounting for age and sex differences. Likewise, ALS prevalence increased by more than double among individuals with T2DM while remaining relatively stable among those without T2DM.

Previous epidemiological studies reported inverse association between T2DM and ALS.^2,3,21^ Mariosa et al. reported in a population-based case-control study of 5108 ALS cases and 25,540 matched population controls during 1991-2010, that non-insulin dependent DM was associated with lower odds of ALS.^2^ In a population-based cohort study in Italy between 1998 to 2014, presence of diabetes at baseline or during follow-up was associated with 70% lower risk of ALS.^21^ Our findings do not necessarily contradict those earlier reports but instead suggest that the relationship between diabetes and ALS may be changing recently. Nor do we infer that a higher body metabolic index (BMI) is generally protective of ALS^22,25^. The reasons for this apparent shift remain uncertain and likely reflect a combination of secular changes in diabetes epidemiology, clinical management, survival, and healthcare utilization.

One potential explanation is the marked evolution in diabetes treatment during the study period. Metformin and insulin prescriptions remained high while sulfonylurea treatment decreased.

Newer glucose-lowering therapies, such as glucagon-like peptide-1 receptor agonists (GLP-1RA) and sodium-glucose cotransporter-2 inhibitors (SGLT-2) treatment significantly increased in this period, coinciding with increasing ALS prevalence among people with T2DM in our cohort. GLP-1RAs in particular are of interest as they are known for their profound weight loss effect.^22^ Several studies have observed the association between lower adiposity and weight loss with increased risk of ALS,^23–26^ and poor outcome among PALS.^10,27,28^ Alternative explanations should also be considered. Improvements in diabetes detection, expansion of participating healthcare systems, changing dietary habits, enhanced electronic health record completeness, and increasing survival among individuals with diabetes could all contribute to increasing co-prevalence independent of any biologic effect.

Our study has limitations. As the current analyses were descriptive and were not designed to evaluate causal relationships between individual medications and ALS risk. Consequently, these temporal trends should not be interpreted as evidence that specific therapies modify ALS susceptibility. Rather, they highlight an important hypothesis that warrants future longitudinal studies incorporating patient-level medication exposure, duration of treatment, metabolic control, and time-to-event analyses. Changes in ALS diagnosis and case ascertainment over time may influence observed prevalence estimates. Although we used validated computable phenotypes and standardized case definitions to maximize specificity, residual ascertainment bias is an inherent limitation of longitudinal EHR-based studies. Lastly, the population included in the study might under-represent younger and healthier population without healthcare utilization and over-representing people with ALS who requires clinical care. Thus, the ALS prevalence in our study shouldn’t be interpreted as true population prevalence of the United States.

## Conclusion and Future Direction

In conclusion, our large-scale, multi-institutional EHR analysis demonstrates that the previously reported inverse relationship between T2DM and ALS has attenuated substantially over the past decade, with T2DM prevalence among PALS now approaching that of the general population and ALS prevalence among individuals with T2DM rising to match the background rate. These converging trends coincide temporally with substantial shifts in the diabetes treatment landscape, including the growing use of GLP-1RAs and their associated weight-loss effects, though our descriptive, population-level design cannot establish a causal link between specific therapies and ALS risk. Rather than resolving the relationship between metabolic disease and motor neuron degeneration, these findings suggest that this relationship is dynamic and may be sensitive to secular changes in treatment practices, diagnostic detection, and survival patterns. Prospective, patient-level studies incorporating detailed medication exposure, timing, and metabolic trajectories will be needed to determine whether these population-level trends reflect a true biological shift in disease risk or are driven by changes in clinical practice and case ascertainment. Given the magnitude and consistency of the trend observed here, this question warrants urgent attention from both the ALS and endocrinology research communities.

## Data Availability

Due to patient privacy protections, institutional data use agreements, and regulatory restrictions, the individual-level data are not publicly available. Access to GROUSE data may be considered for qualified researchers upon reasonable request, subject to institutional approvals, applicable data governance policies, and execution of appropriate data use agreements.

## Acknowledgments

This work utilized EHR data and research infrastructure from the GPC, a consortium within the National Patient-Centered Clinical Research Network (PCORnet®). We acknowledge the contributions of GPC participating institutions and their research and informatics teams. The data and infrastructure were originally supported by a Patient-Centered Outcomes Research Institute® (PCORI®) Program Award (#RI-MISSOURI-01-PS8).

---

**Supplementary Table 1.**
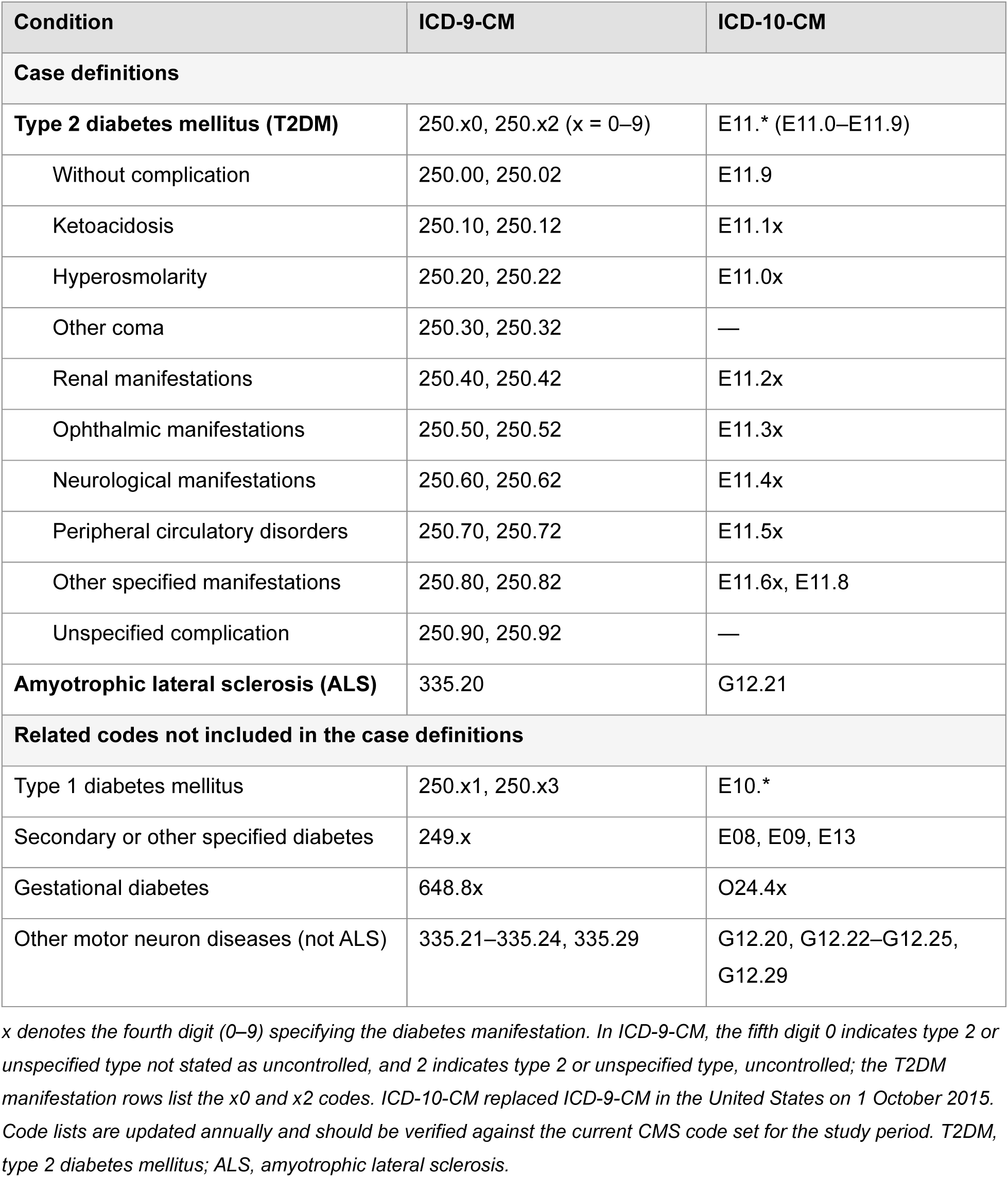
ICD-9-CM and ICD-10-CM diagnosis codes for T2DM and ALS.

## Notes

### Competing Interest Statement

The authors have declared no competing interest.

### Author Declarations

The use of data from the GROUSE platform for this study was reviewed and approved by the Institutional Review Board (IRB) of the University of Missouri (IRB# 2057285). The study was determined to be exempt from further IRB review and was granted a full waiver of HIPAA authorization.

