## Supplemental Table 1 for "The prevalence of type 2 diabetes mellitus among people with amyotrophic lateral sclerosis is increasing in the United States"

**Supplementary Table 1. ICD-9-CM and ICD-10-CM diagnosis codes for T2DM and ALS**

| Condition | ICD-9-CM | ICD-10-CM |
|---|---|---|
| **Case definitions** | | |
| **Type 2 diabetes mellitus (T2DM)** | 250.x0, 250.x2 (x = 0–9) | E11.* (E11.0–E11.9) |
| Without complication | 250.00, 250.02 | E11.9 |
| Ketoacidosis | 250.10, 250.12 | E11.1x |
| Hyperosmolarity | 250.20, 250.22 | E11.0x |
| Other coma | 250.30, 250.32 | — |
| Renal manifestations | 250.40, 250.42 | E11.2x |
| Ophthalmic manifestations | 250.50, 250.52 | E11.3x |
| Neurological manifestations | 250.60, 250.62 | E11.4x |
| Peripheral circulatory disorders | 250.70, 250.72 | E11.5x |
| Other specified manifestations | 250.80, 250.82 | E11.6x, E11.8 |
| Unspecified complication | 250.90, 250.92 | — |
| **Amyotrophic lateral sclerosis (ALS)** | 335.20 | G12.21 |
| **Related codes not included in the case definitions** | | |
| Type 1 diabetes mellitus | 250.x1, 250.x3 | E10.* |
| Secondary or other specified diabetes | 249.x | E08, E09, E13 |
| Gestational diabetes | 648.8x | O24.4x |
| Other motor neuron diseases (not ALS) | 335.21–335.24, 335.29 | G12.20, G12.22–G12.25, G12.29 |

*x denotes the fourth digit (0–9) specifying the diabetes manifestation. In ICD-9-CM, the fifth digit 0 indicates type 2 or unspecified type not stated as uncontrolled, and 2 indicates type 2 or unspecified type, uncontrolled; the T2DM manifestation rows list the x0 and x2 codes. ICD-10-CM replaced ICD-9-CM in the United States on 1 October 2015. Code lists are updated annually and should be verified against the current CMS code set for the study period. T2DM, type 2 diabetes mellitus; ALS, amyotrophic lateral sclerosis.*
